# Are there causal relationships between ADHD and BMI? Evidence from multiple genetically informed designs

**DOI:** 10.1101/2020.04.16.20067918

**Authors:** Chao-Yu Liu, Tabea Schoeler, Neil M Davies, Hugo Peyre, Kai-Xiang Lim, Edward D Barker, Clare Llewellyn, Frank Dudbridge, Jean-Baptiste Pingault

## Abstract

**Background:** Attention-deficit/hyperactivity disorder (ADHD) and Body Mass Index (BMI) are associated. However, it remains unclear whether this association reflects causal relationships in either direction, or confounding. Here, we implemented genetically informed methods to examine bidirectional causality and potential confounding.

**Methods:** Three genetically informed methods were employed: (1) cross-lagged twin-differences analysis to assess bidirectional effects of ADHD symptoms and BMI at ages 8, 12, 14 and 16 years in 2,386 pairs of monozygotic twins from the Twins Early Development Study (TEDS), (2) within- and between-family ADHD and BMI polygenic score (PS) analysis in 3,320 pairs of dizygotic TEDS twins and (3) two-sample bidirectional Mendelian randomization (MR) using summary statistics from Genome-Wide Association Studies (GWAS) on ADHD (N=55,374) and BMI (N=806,834).

**Results:** Mixed results were obtained across the three methods. Twin-difference analyses provided little support for cross-lagged associations between ADHD symptoms and BMI over time. PS analyses were consistent with bidirectional relationships between ADHD and BMI with plausible time-varying effects from childhood to adolescence. MR findings were also consistent with bidirectional causal effects between ADHD and BMI. Multivariable MR suggested the presence of substantial confounding in bidirectional relationships.

**Conclusions:** The three methods converged to highlight multiple sources of confounding in the association between ADHD and BMI. PS and MR analyses suggested plausible causal relationships in both directions. Possible explanations for mixed causal findings across methods are discussed.

**Key messages:**

- Within-family polygenic score and Mendelian randomization analyses were consistent with bidirectional causal effects between ADHD and BMI.
- Findings from different genetically informed methods suggested that multiple sources of confounding are at play, including genetic and shared environmental confounding, population stratification, assortative mating and dynastic effects.
- The ADHD polygenic score increasingly associated with BMI phenotype from childhood to adolescence, suggesting an increasing role of ADHD in the aetiology of BMI across the development. Conversely, BMI polygenic score association with ADHD phenotypes tended to decrease across the development.
- Addressing mixed evidence will require increased sample sizes to implement novel methods such as within-family MR.

## Introduction

Two meta-analyses confirm the association between Attention-Deficit Hyperactive Disorder (ADHD) and overweight/obesity with pooled odds ratios ranging from 1.22 to 1.27 (1, 2). Such positive associations between ADHD and obesity are more established in late adolescence and adulthood, while findings in children are frequently mixed (1-3). Prospective studies indicate that the relationships between ADHD and obesity may be bidirectional. Evidence showing that ADHD symptoms precede overweight and obesity (4, 5) and that overweight and obesity lead to manifestations of ADHD (6) (7) can all be found in the literature. Indeed, the behavioural characteristics of ADHD (e.g. lack of planning, poor impulse control) may increase the risk of overeating and abnormal eating behaviours, leading to weight gain and obesity (8). Conversely, obesity-related neurocognitive dysfunction (9), brain structural abnormality (10), inflammation (11) and sleep disruption (12) may cause impairment in attention and inhibitory control.

In addition to plausible bidirectional causal relationships, it is also possible that the association between ADHD and obesity arises, totally or in part, from confounding. Previous investigations showed that the association between ADHD and overweight/obesity significantly reduced after adjusting for socioeconomic and lifestyle factors (13). Furthermore, family aggregation of overweight/obesity and ADHD was better explained by shared environmental factors than by direct effects between the two phenotypes (14). In addition to environmental confounding, genetic confounding can also play a role. Although positive genetic correlations (ranging from r = 0.21 to 0.26) and polygenic score associations between ADHD and BMI have been identified (15-18), such genetic relationships may reflect a causal cascade (mediated pleiotropy, also called vertical pleiotropy), or genetic confounding (unmediated pleiotropy, also called horizontal pleiotropy), or a combination of both (19, 20). Therefore, a genetic association can arise in the absence of causal relationships.

To better differentiate between causality and confounding, a range of genetically informed methods for causal inference can be implemented (21). Importantly, using several such approaches to address the same question remains necessary to triangulate evidence, as results from genetically informed methods can still be biased or confounded. Triangulating evidence from different methods can not only identify potential sources of bias, but also strengthen causal inference if different methods point to the same conclusion (22). For example, a recent study used genetic variants associated with ADHD and BMI as instrumental variables in Mendelian randomization (MR) analyses and detected an effect from higher BMI to higher ADHD liability (23) but not in the reverse direction. These findings were at odds with previous observational reports (1). Nevertheless, MR is susceptible to unmeasured confounding such as dynastic effects (i.e. when associations between offspring genetic variants and offspring phenotypes also capture environmentally mediated parental genetic effects) and population stratification (24), it is important to validate these findings with methods controlling for aforementioned confounding, such as family-based genetically informed designs (e.g. within-family analysis) (21).

In this study, we implemented three different genetically informed methods to investigate the nature of the relationship between ADHD and BMI: 1) cross-lagged twin-differences analysis in MZ twins to examine the causal effects across childhood and adolescence while controlling for shared genetic and shared environmental confounding, 2) between- and within-family polygenic score analyses, to account for dynastic effects, population stratification and assortative mating shared between siblings, and 3) Mendelian randomization and corollary sensitivity analyses to account for unmediated pleiotropy. We aimed to investigate to what extent the association between ADHD and BMI is attributable to causal relationships and/or to other factors.

## Methods

The description, strengths and assumptions of the three genetically informed methods, namely (1) twin-differences, (2) polygenic scoring and (3) bidirectional Mendelian randomization are outlined in Table 1. The following sections detail the implementation of those methods in the current study.

**Table 1.** Description of the three inference methods employed to study the relationship between ADHD and BMI

| Approach | General description | Rationale and aims for application in current study | Main assumptions |
| --- | --- | --- | --- |
| 1. Twin-differences analysis | Twin-differences analysis capitalises on the twins' characteristics to control for potential confounding due to genetic and shared environmental effects. If one twin differs from the co-twin in one trait (e.g. BMI) also differs in the outcome of interest (e.g. ADHD), then it suggests effects are independent from genetic and shared environment influences. | **Twin-differences analysis in MZ twins**<br>• MZ twins provide a stringent control for genetic and shared environmental confounding.<br>• The longitudinal design assesses whether differences in one trait associate with subsequent differences in the other trait.<br>• Twin-differences analyses can inform if the effects are due to non-shared environment or confounding. | MZ twins share 100% of their genetic material (although small divergences from this general rule can be observed). MZ twin-differences analysis assumes that there is no unobserved non-shared environmental confounding. Plausible observed confounders (here birth weight) can be adjusted for. |
| 2. Polygenic score (PS) analysis | A PS for a specific trait per individual is derived by computing the sum of the trait-associated alleles weighted by their relative effect sizes as reported in the genome-wide association summary statistics. | **Between- and within-family PS association in DZ twins**<br>• A PS indexes an individual's genetic liability to a particular trait, thus can be used as a proxy of that trait in cross-trait analyses. Estimates obtained from a PS association can be mathematically identical to causal estimates obtained from the IVW estimator in Mendelian randomization (MR), as long as the same genetic variants and GWAS estimates are used in both methods (see the MR<br>section below). PS associations provide an initial indication of a possible causal relationship.<br>• The within-family design controls for dynastic effects<sup>1</sup>, population stratification<sup>2</sup> and assortative mating<sup>3</sup>, which can bias PS association findings.<br>• Implementing within-family polygenic score association as performed in the present study addresses confounding of dynastic effects that are common to the PS association and the MR approach (see assumption column). | • The main assumption for the use of PS in the context of causal inference is the absence of pleiotropy. However, this assumption is most unlikely to hold. |
| 3. Bidirectional Mendelian randomization analysis (MR) | MR analysis builds on Mendel's Laws of Inheritance, and utilises genetic variants associated with an exposure (e.g. SNPs associated with BMI) as instrumental variable (IV) to examine the causal effect of this exposure on an outcome. | **Bi-directional two-sample MR**<br>• Evaluate reciprocal relationships between ADHD and BMI.<br>**IVW-based method**<br>• The IVW regression estimates the causal effect by computing the slope of the weighted regression of instrument-outcome associations on instrument-exposure associations, with the intercept constrained to zero.<br>**MR-Egger regression**<br>• The intercept estimate can be interpreted as the average unbalanced horizontal pleiotropic effects across | • Three core instrumental assumptions regarding the IV should be met for all MR analyses:<br>○ Relevance assumption: the genetic variants are associated with the exposure.<br>○ Independence assumption: association between the genetic variants and the outcome is not confounded by unmeasured factors.<br>○ Exclusion restriction: genetic variants only affect the outcome through their effect on the exposure. Unmediated pleiotropy (also called |
|  |  | the IVs.<br>**Weighted-median method**<br>Provides a valid causal estimate if the IVs that represent 50% of the weight in the estimate are valid.<br>**Weighted-mode method**<br>Provides a valid causal estimate if the IVs that represent the weighted mode (i.e. most common) represent the true causal effect. It is robust to unbalanced horizontal pleiotropy and is unbiased when the majority of the IVs are invalid.<br>**Robust adjusted profile score (MR-RAPS)**<br>Attributes different weights to the IVs according to their associative strengths to minimise the weak instrument bias. It enables two-sample MR analysis to include more weak IVs, which is not recommended with other methods for MR, and increases efficiency of MR studies.<br>**Multivariable MR analysis**<br>Provides effect estimates of multiple exposures on one outcome, thereby explicitly controlling for suspected sources of pleiotropy. Here, we<br>implement multivariable MR by modelling genetic variants associated with educational attainment to test whether genetic nurture effects associated with educational attainment may explain away some of the reciprocal effects between ADHD and BMI. | horizontal pleiotropy<sup>4</sup>) violates this assumption.<br>**IVW-estimator** only accounts for balanced pleiotropy<sup>5</sup>, i.e. when average pleiotropic effect equals to zero, simply creating additional heterogeneity without modifying the point estimate. Conversely, **MR-Egger** provides an estimate of unbalanced horizontal pleiotropy and can yield a correct estimate even if all instruments are invalid. But **MR-Egger** has low power.<br>**The weighted-median method** can provide accurate causal estimates when > 50% of the instrument are valid and is more powerful than **MR-Egger**.<br>**The weighted-mode method** provides a valid causal estimate if the largest subset of IVs with the same ratio of effect is formed by valid instruments.<br>**MR-RAPS** can be employed to correct for the weak instrument bias and also accounts for balanced pleiotropy.<br>**Multivariable MR analysis** provides accurate causal effects if sources of pleiotropy are effectively captured and explicitly modelled. |

Note: <sup>1</sup> Dynastic effect: non-transmitted alleles from parents influence offspring phenotype via the correlation between the environment parents create for their children and the non-transmitted gene. <sup>2</sup>Population stratification: the presence of a systematic difference in allele frequencies between subgroups in a population. It mainly results from non-random mating due to geographic separation and low rates of migration. <sup>3</sup>Assortative mating: individuals with similar phenotype mate with one another more frequently than that would be expected at random. For example, married couples often share similar level of education or socioeconomic background. <sup>4</sup>Horizontal pleiotropy, when one genetic variant has independent effect on multiple traits. <sup>5</sup>Balanced pleiotropy, on average the pleiotropic effects on the outcome equal to zero (hence balanced) and is independent of the pleiotropic effects on the exposure. When the average pleiotropic effect is positive or negative, the horizontal pleiotropy is unbalanced.

### I. Twin-differences analyses

#### Study Sample

Data were drawn from the Twins Early Development Study (TEDS), a cohort of twins born between 1994 and 1996 in England and Wales. Additional details on the TEDS sample can be found elsewhere (25). Four waves of data collection including data on BMI and ADHD symptoms were analysed (ages 8, 12, 14 and 16 years). In this study, we included twin pairs with complete information on zygosity and ratings of ADHD symptoms and BMI across at least one of the four assessment waves. Because only a subset of twin pairs has genotyping data, zygosity information was derived from an algorithm based on parent-reported phenotypic similarity. However, if a DNA zygosity test is available, the information was used as best estimate for a same-sex twin pair. The zygosity assignment by the algorithm has 95.2% concordance with DNA findings in the TEDS sample (25, 26). Our final study sample comprised 6,655 twin pairs (48% males), including 2386 monozygotic twin (MZ) pairs and 4,269 dizygotic twin (DZ) pairs.

Ethical approval for the Twins Early Development Study (TEDS) was granted by King’s College London’s ethics committee for the Institute of Psychiatry, Psychology and Neuroscience. Written informed consent was obtained from parents before data collection.

#### Phenotypic measures for ADHD symptoms and BMI

ADHD symptom scores were derived from parents’ ratings on the Conners’ Parent Rating Scale-Revised (CPRS-R) at ages 8, 12, 14 and 16 years. The CPRS-R consists of 18 items tapping inattentive and hyperactive/impulsive symptoms of ADHD (27). Evidence suggests that the CPRS-R is a reliable measure to assess ADHD and the diagnostic effectiveness of the CPRS-R has been demonstrated across different settings (28). All items were rated on a 0-3 Likert scale, with 0 as ‘not at all’ and 3 as ‘very much true’. A sum score ranging from 0 to 54 was calculated from the 18 items of the CPRS-R, indexing the levels of ADHD symptoms of each individual. The information was used in phenotypic, twin-differences, and polygenic score analyses. Standardised Cronbach alphas for the CPRS-R total scale ranged between 0.91 and 0.92 across the 4 waves of data collection. BMI of the twins was obtained using parent-reported data on height (meter) and weight (kilogram) at age 8, and child-reported data at ages 12, 14 and 16 years. BMI values were converted to age- and sex-adjusted standardised deviation scores (SDS) using the LMS method (29) based on the British 1990 growth reference (30).

#### Analyses

All statistical analyses were conducted using R [version 3.5.2 (31)].

We used structural equation modelling [R package Lavaan (32)] to construct analysis with cross-lagged design (33) and derive the following estimates:

- Cross-lagged phenotypic associations between earlier measures of ADHD symptoms (BMI SDS) and later measures of BMI SDS (ADHD symptoms), adjusted for observed confounds (sex, age, birth weight and parental education) on 6,655 unrelated individuals (one twin selected at random from each twin pair) across childhood and adolescence.
- A cross-lagged twin-differences model on MZ twins (2,386 pairs) to examine the relationships between differences in earlier measures of ADHD symptoms (BMI SDS) and later measures of BMI SDS (ADHD symptoms) across childhood and adolescence. Sex, age and parental education were adjusted by design and a twin- differences score in birth weight was included as a covariate (details in Table 1).

To account for data non-normality, 95% confidence intervals (CI) were obtained using bootstrapping with 10,000 repetitions.

### II. Polygenic Score analyses

#### Study Sample

A subsample of 3,320 dizygotic (DZ) twin pairs from the TEDS that had complete phenotype data and passed the genotyping quality control procedures were included in the polygenic score analyses (details in (34)). Observations of each twin and the co-twin were entered in the multilevel model to conduct a family-based design analysis. Such approach provides association estimates of the polygenic scores on the phenotypic traits unbiased by dynastic effects, population stratification and assortative mating (definitions and details in Table 1) (35).

#### Genotypic data and polygenic scores

The ADHD and BMI polygenic scores (PSs) for the TEDS participants were generated in the software LDpred (36), using the TEDS genotype data and the summary statistics from genome-wide association studies (GWAS) on ADHD (N=55,374) (15) and BMI (N=806,834) (17). The PSs for each participant were computed based on a fraction of causal markers of 1 in LDpred (see (37) for details on PS generation). LDpred method accounts for linkage disequilibrium (LD) between SNPs. To facilitate interpretability, the PSs were z-standardised with a mean=0 and standard deviation=1. The ADHD PS explained 1.1 to 1.8% of the total variance in ADHD symptoms, while the BMI PS explained 2.7 to 11.7 % of the total variance in BMI SDS (Supplementary Table 1).

#### Analyses

Multilevel models using the package “lme4” (38) were implemented to test the associations between ADHD/BMI PSs and their opposite phenotype. Because twins are nested within families, the model clustered standard errors by family and allowed for within-family correlations. ADHD ratings, BMI SDS, and the polygenic scores for ADHD and BMI were residualised on age, sex, and the first 10 principal components prior to estimating the multilevel model.

For each multilevel model, we estimated the associations between the ADHD PS and BMI phenotypes and the BMI PS and ADHD ratings with:

- The family mean PS (i.e. the averaged PS across the two twins): this estimates the between-family association.
- The difference between the individual PS and the family mean PS: this estimates the within-family association.

We then computed the following differences in estimates from the multilevel models:

- Differences of the within-family and the between-family estimates (the within-family minus the between-family estimates): to examine estimate change due to the three aforementioned sources of bias.
- Differences of the within-family estimates between ages 16 and 8 years (estimate at age 16 minus estimate at age 8): to examine whether effects varied across development.

Differences were tested against the null using 95% bootstrap percentile intervals based on random sampling with replacement of DZ twin pairs (10,000 draws).

It has been shown that the between-family estimates can be confounded by dynastic effects related to parental education (37). We therefore included parental education (standardized average of maternal and paternal highest educational level collected at first contact) in our model to test if it changed the between-family estimates. As above, we tested the following differences:

- Differences of the within-family and the between-family estimates in the model, this time including parental education as a covariate.
- Differences of the between-family estimate before and after adjusting for parental education.

### III. Bidirectional two-sample Mendelian Randomization (MR) analyses of ADHD and BMI

#### Study Samples

MR is a causal inference method that uses genetic variants associated with an exposure (e.g. SNPs associated with BMI) as instrumental variables to estimate the effect of this exposure on an outcome (Table 1 for details). We selected SNPs below the genome-wide significance p-value threshold of p<5e-8 (i.e. 5×10^−8^) for BMI (N_SNPs_=546) and SNPs below the suggestive p-value threshold of p<5e-5 for ADHD (N_SNPs_= 190) with clumping to ensure independence between SNPs (clumping r^2^ cut off =0.001 and clumping window=10,000kb) from recently published genome-wide association (GWA) summary data on BMI (N=806,834, (17)) and ADHD (N=55,374, (15)).

We also performed MR analyses with SNPs selected at the p-value threshold of p<5e-8 from the ADHD GWA summary data to check for the convergence in findings with analyses at the p-value threshold of p<5e-5.

#### Analyses

We used the TwoSampleMR package (39) for MR analyses. We considered ADHD and BMI as exposures in turn to evaluate bidirectional effects. Effect estimates from individual SNPs were combined using random-effects inverse-variance weighted (MR-IVW) regression as the primary analysis.

To interrogate possible violations of key MR assumptions and to deal with a potential weak instrument bias, we conducted a number of sensitivity analyses, including (see details in Table 1):

- MR-Egger analysis
- Weighted median analysis
- Weighted mode analysis
- Robust adjusted profile score (MR-RAPS) analysis
- Multivariable MR using the IVW estimator to estimate the direct effects of BMI and ADHD after controlling for confounding associated with educational attainment

Previous findings showed that polygenic scores for educational attainment were associated with large dynastic effects on a number of traits including BMI (40). To examine whether controlling for educational attainment influences some of the reciprocal effects between ADHD and BMI, we compared the IVW MR estimator with the IVW multivariable MR estimator using a test that accounts for the covariance between the two estimates (41).

## Results

### I. Twin-differences analyses

Sample baseline characteristics are shown in Supplementary Table 2. Figure 1 and Supplementary Table 3 present the autoregressive and cross-lagged phenotypic associations between ADHD symptoms and BMI SDS from ages 8 to 16 years, adjusted for age, sex, birth weight and parental education in unrelated individuals. Evidence indicated that ADHD symptoms were positively associated with subsequent BMI across the follow up (ß=0.028, 95% bootstrap CI=0.022,0.034, ß=0.033, 95% bootstrap CI=0. 0.027,0.039 and ß=0.045, 95% bootstrap CI=0.037,0.053 respectively) (Supplementary Table 3). Conversely, evidence supported that BMI at age 14 was negatively associated with ADHD symptoms at age 16 years (ß=-0.010, 95% bootstrap CI=-0.018,-0.002).

**Figure 1.**
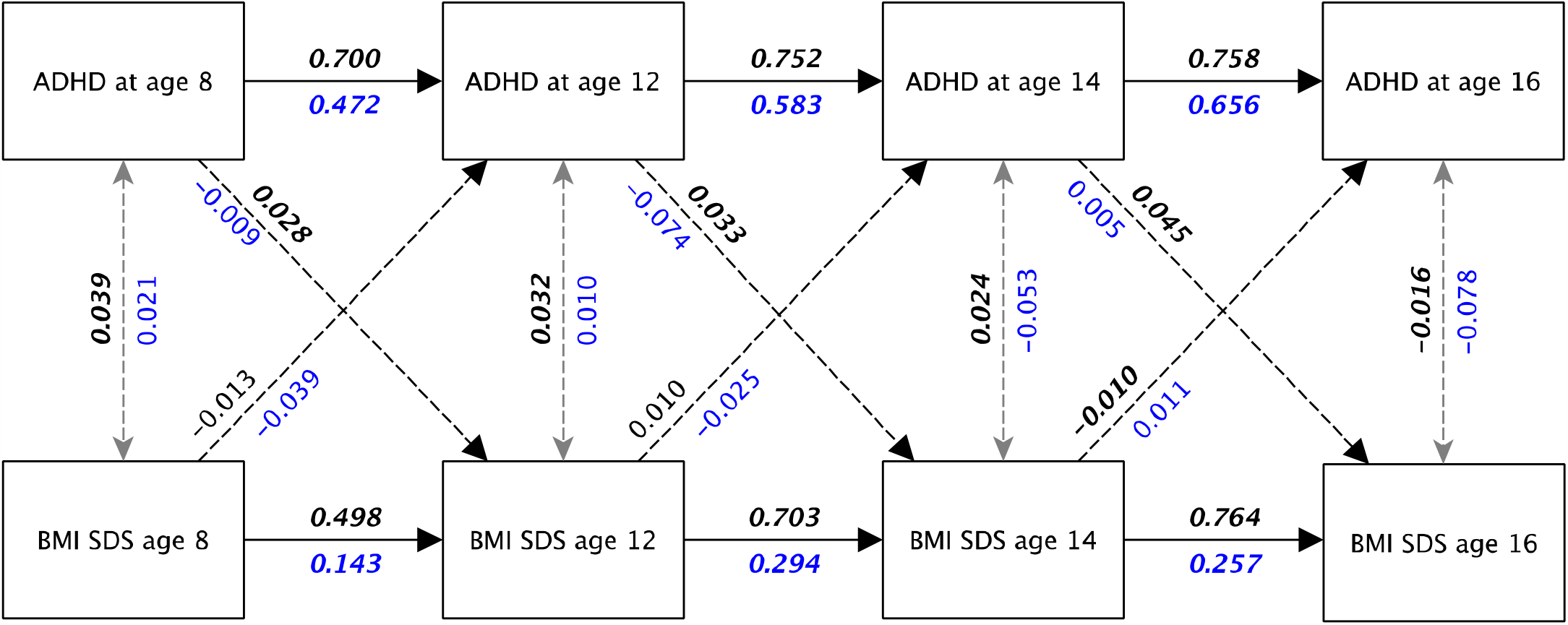
Cross-lagged phenotypic relationships between ADHD symptoms and BMI SDS Note: Standardised estimates of phenotypic relationships (i.e. correlations) adjusted for age, sex, birth weight and parental education are shown in black (above the lines); estimates from twin-differences analyses in MZ twins are in blue (below the lines); estimates with 95% bootstrap percentile intervals not including 0 are displayed in ***italic bold***. Details of 95% bootstrap percentile intervals can be found in Supplementary Table 3 and Supplementary Table 4.

Results of the twin-differences analyses on MZ twins are shown in Figure 1 and Supplementary Table 4. MZ twin-differences analyses provided little support that differences in earlier measures of ADHD symptoms were associated with subsequent changes in BMI (ß=-0.009, −0.074 and 0.005, all 95% bootstrap CI included 0) and vice versa (ß=-0.040, - 0.025 and 0.011, all 95% bootstrap CI included 0). Means and distribution of the differences in standardised ADHD symptom ratings and BMI SDS in MZ twins at ages 8, 12, 14 and 16 are shown in Supplementary Figure 1.

### II. Polygenic score analyses

#### ADHD Polygenic score to BMI SDS

The associations between the ADHD PS and phenotypic BMI from the multilevel model are displayed in Figure 2 and Supplementary Table 5. Findings indicated positive between-family associations at ages 12, 14 and 16 years (e.g. at age 12 years, Beta=0.057, 95%CI=0.016,0.098, i.e. one SD unit increase in the ADHD PS was associated with 0.057 SD unit increase in BMI SDS). The within-family estimates provided evidence that a one SD unit increase in the individual ADHD PS from the family mean was associated with a 0.128 (95%CI 0.025 to 0.228) SD unit increase in BMI SDS at age 16 years. There was little evidence that the ADHD PS associated with BMI within families at earlier ages. There was little evidence that the between-family and the within-family estimates differed (Δ=-0.041 to 0.040, bootstrap 95% CIs all across 0) (Supplementary Table 5). Overall, the within-family associations increased from age 8 to age 16 years (Δ=0.088, 95% bootstrap CI 0.038,0.214).

**Figure 2.**
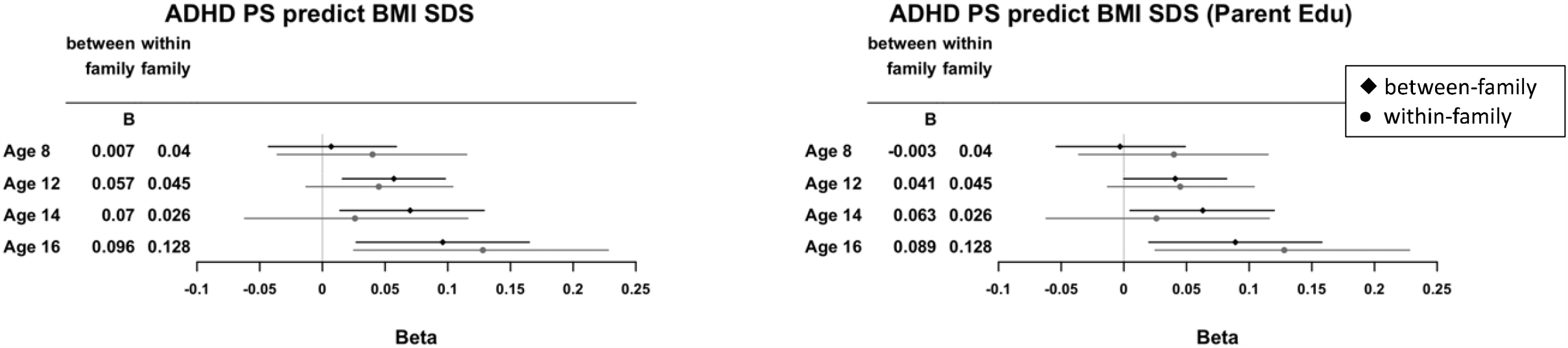
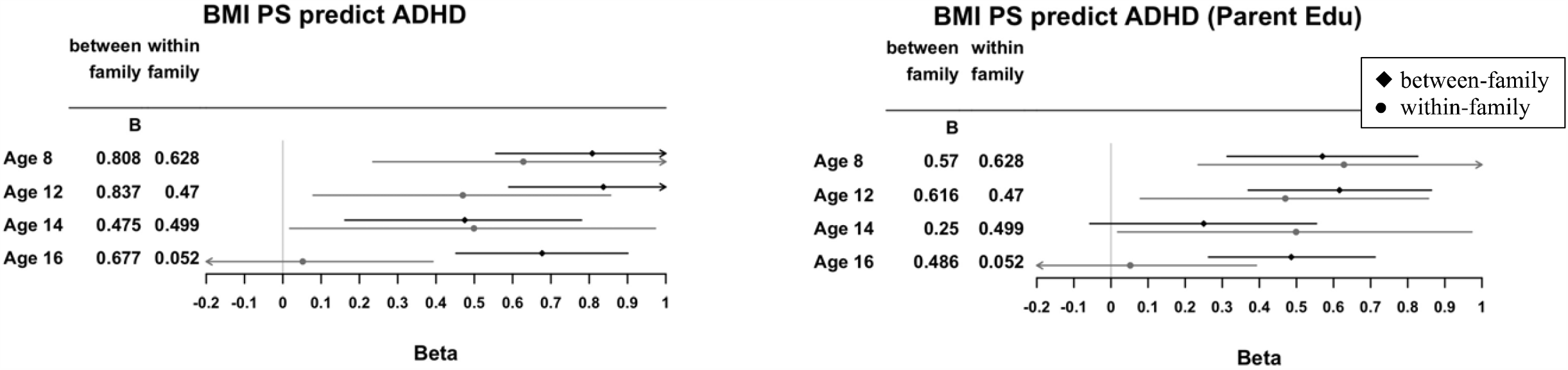
Between- and within-family polygenic score associations Note: PS, standardised polygenic score; B/Beta, unstandardised regression estimates.

After including parental education level as a covariate, the between-family associations were attenuated at ages 8, 12 and 14 years (Δ=-0.010, −0.016 and −0.009, bootstrap 95% CIs do not include 0), while the within-family associations remained unchanged. In sum, the effects of the ADHD PS on phenotypic BMI increased from childhood to adolescence and were confounded by parental education (Figure 2 and Supplementary Table 5).

#### BMI polygenic score to ADHD symptoms

The associations between the BMI PS and ADHD symptoms are shown in Figure 2 and Supplementary Table 6. The BMI PS was positively associated with ADHD symptoms between families across the four time points (e.g. at age 8 years, Beta=0.808, 95%CI=0.556,1.061, i.e. one SD unit increase in BMI PS was associated with 0.808-point increase in the score of ADHD symptoms at age 8 years), and there was evidence of within-family associations at ages 8, 12 and 14 years (e.g. at age 8 years, one SD unit increase in the individual BMI PS from the family mean was associated with a 0.628-point (95%CI 0.235,1.022) increase in the score of ADHD symptoms). Evidence showed that the within-family association was smaller than the between-family association at age 16 years (Δ=-0.626, bootstrap 95% CI -1.029,-0.217) (Supplementary Table 6) but not at earlier ages. In contrast to the ADHD PS, the effects of the BMI PS on ADHD symptoms decreased from age 8 to age 16 years (Δ=-0.538, 95% bootstrap CI −0.988,-0.094).

After including parental education level as a covariate, the between-family associations were attenuated at ages 8, 12 and 16 years (Δ=-0.238, −0.220 and −0.434, bootstrap 95% CIs do not cross 0) (Supplementary Table 6). In sum, the effects of the BMI PS on ADHD symptoms decreased from childhood to adolescence and were confounded by parental education.

### III. Bidirectional two-sample Mendelian Randomization (TSMR)

#### Effect of ADHD liability on BMI

After harmonisation to exclude palindromic SNPs and variants that did not share the same allele pair between the exposure and the outcome GWAS, 131 of the 190 SNPs selected at the threshold of p<5e-5 were used as genetic instruments for ADHD. Table 2 and Figure 3 show the effect of the liability to ADHD on BMI estimated by different MR methods. The estimates suggested that a unit increase in odds ratio of ADHD lead to 0.028 SD unit increase in BMI (IVW ß =0.028, 95%CI 0.015-0.040, p<0.001). The sensitivity analyses using weighted median method and MR-RAPS were consistent with the IVW results. The weighted mode method and the MR Egger estimates were imprecise (Table 2). MR-Egger intercept provided little evidence of unbalanced horizontal pleiotropy (intercept=0.002, 95%CI −0.002- 0.005, p=0.330). The sensitivity analyses performed with the SNPs below the genome-wide significance of p<5e-8 showed that one unit increase in odds ratio of ADHD lead to 0.064 SD unit increase in BMI (IVW ß =0.064, 95%CI 0.004-0.125, p=0.036). The weighted median method was consistent with the IVW, but the weighted mode method and MR Egger provided little support for an effect (Supplementary Table 7).

**Table 2.** Mendelian randomization: effects between ADHD and BMI

| Exposure | Method | Univariable MR |  |  |  |  | Multivariable MR |  |  |  |  |
| --- | --- | --- | --- | --- | --- | --- | --- | --- | --- | --- | --- |
| | | nSNP | $\beta$ | Lower<br>95% CI | Upper<br>95% CI | *p*-value | nSNP | $\beta$ | Lower<br>95% CI | Upper<br>95% CI | *p*-value |
| ADHD | IVW | 131 | 0.028 | 0.015 | 0.040 | **<0.001** | 130 | 0.019 | 0.005 | 0.033 | **0.010** |
|  | MR RAPS |  | 0.020 | 0.010 | 0.031 | **<0.001** |  |  |  |  |  |
|  | Weighted median |  | 0.013 | 0.004 | 0.023 | **0.005** |  |  |  |  |  |
|  | Weighted mode |  | 0.002 | -0.024 | 0.027 | 0.905 |  |  |  |  |  |
|  | MR Egger |  | 0.008 | -0.034 | 0.049 | 0.713 |  |  |  |  |  |
|  | MR Egger intercept |  | 0.002 | -0.002 | 0.005 | 0.330 |  |  |  |  |  |
| BMI | IVW | 463 | 1.923* | 1.715 | 2.157 | **<0.001** | 459 | 1.587* | 1.395 | 1.806 | **<0.001** |
|  | MR RAPS |  | 1.910* | 1.699 | 2.147 | **<0.001** |  |  |  |  |  |
|  | Weighted median |  | 1.973* | 1.710 | 2.276 | **<0.001** |  |  |  |  |  |
|  | Weighted mode |  | 2.353* | 1.570 | 3.527 | **<0.001** |  |  |  |  |  |
|  | MR Egger |  | 1.679* | 1.228 | 2.297 | **0.001** |  |  |  |  |  |
|  | MR Egger intercept |  | 0.002 | -0.003 | 0.007 | 0.362 |  |  |  |  |  |

Note: \* is odds ratio (OR).

**Figure 3.**
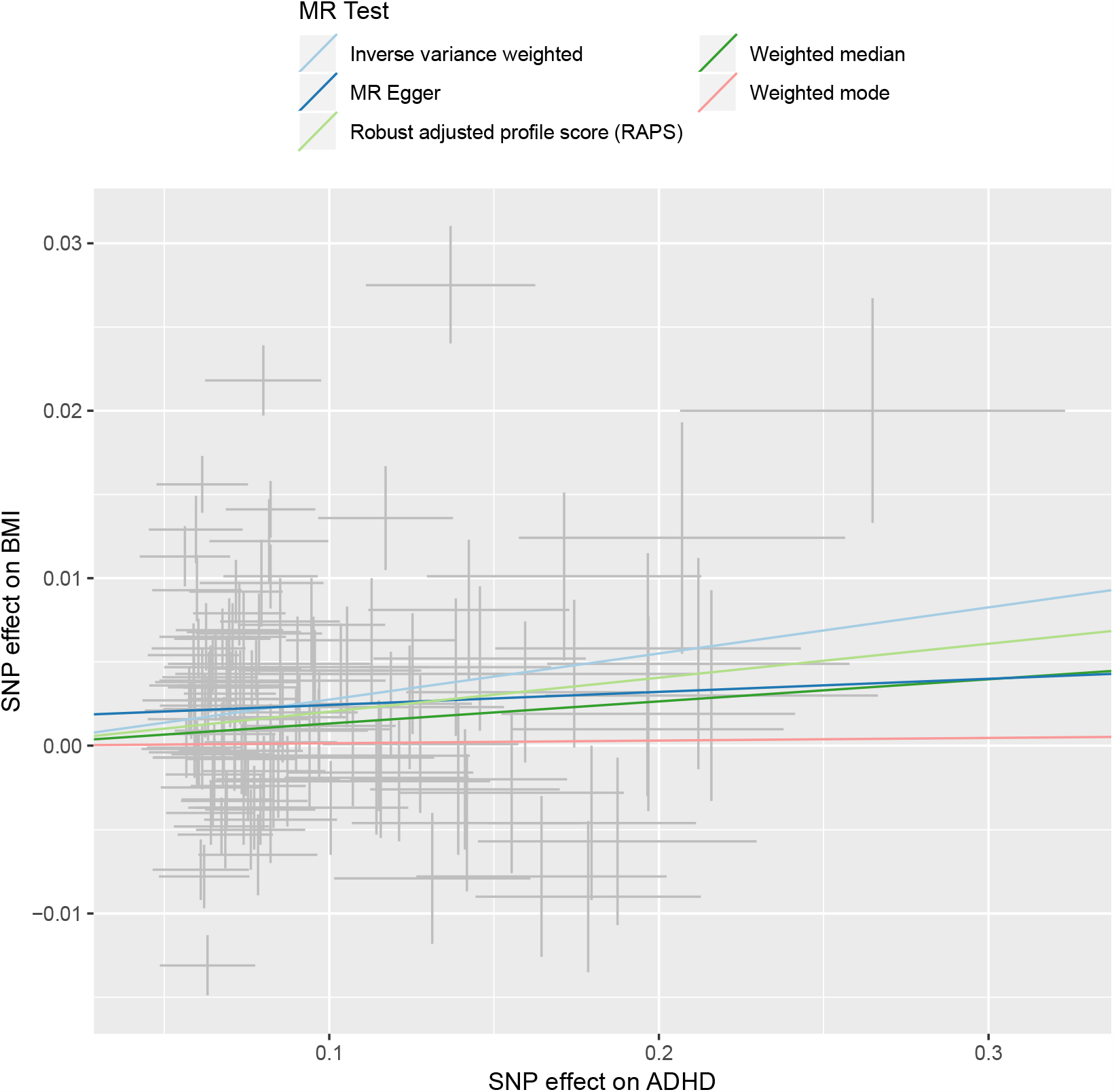
Mendelian randomization: effects of ADHD on BMI

The multivariable IVW estimate of the effect of liability to ADHD on BMI controlling for effects associated with education, was smaller than the univariable IVW estimate (multivariable MR ß =0.019, 95%CI 0.005-0.033, p=0.010), as found in the test of differences (t(127)=2.362, p=0.020).

#### Effect of BMI on the liability to ADHD

After harmonisation, 463 of the 546 SNPs selected from the BMI GWAS below the genome-wide significance of p<5e-8 were used as genetic instruments for BMI. As shown in Figure 4, different MR estimators consistently provided evidence that higher BMI increases liability to ADHD. The estimate of MR-IVW (IVW OR=1.923, 95%CI 1.715-2.157, p<0.001) suggested that a one SD unit increase in BMI nearly doubled the odds of ADHD. MR-Egger intercept provided little evidence of unbalanced horizontal pleiotropy (intercept=0.002, 95%CI −0.003- 0.007, p=0.362) (Table 2).

**Figure 4.**
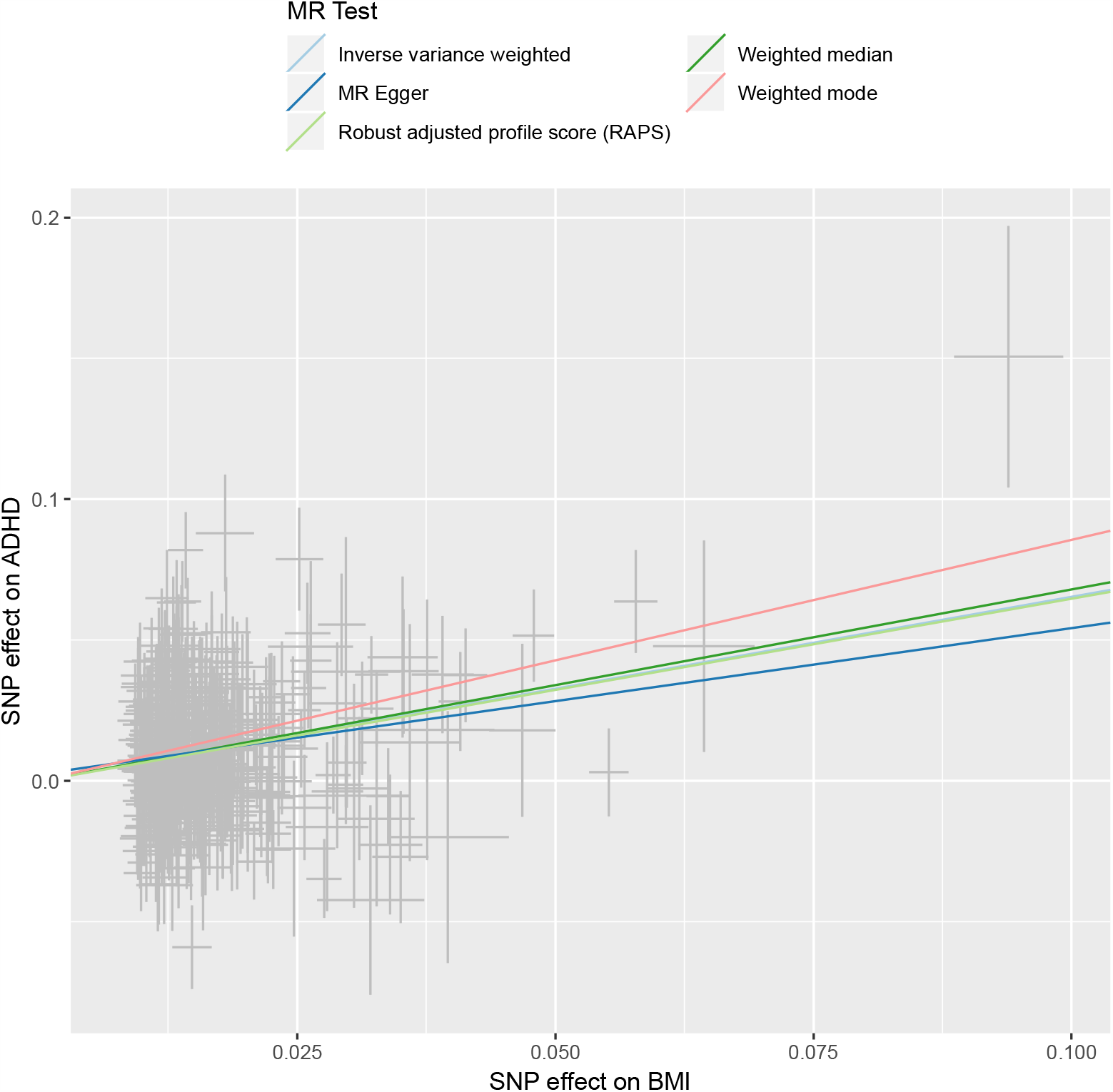
Mendelian randomization: effects of BMI on ADHD

The MVMR estimate also provided evidence that BMI had an effect on liability to ADHD that was independent of effects associated with education (multivariable MR-IVW OR=1.587, 95%CI 1.395-7.583, p<0.001), although the effect was smaller as found in the test of difference (t(456)=5.636, p<0.001).

## Discussion

The current study used three different genetically informed methods (MZ twin-differences, polygenic score and Mendelian randomization analyses) to test the nature of the relationship between ADHD and BMI. Triangulation of evidence from the three methods indicated multiple sources of confounding such as population stratification, assortative mating and dynastic effects underlying the relationship between ADHD and BMI. Findings suggested that some of the effects between ADHD and BMI may be confounded by effects associated with education. Possible explanations for mixed causal findings across methods are discussed.

### The relationship between ADHD and BMI

#### Unidirectional or bidirectional?

In cross-lagged phenotypic analyses on unrelated individuals, we found that ADHD symptoms predicted higher BMI at later ages but not the reverse. However, findings from cross-lagged MZ twin differences provided little support for an effect of ADHD on later BMI and vice versa. In contrast, polygenic score and MR analyses were consistent with the presence of bidirectional relationships between ADHD and BMI. There was more consistent evidence supporting the effect of BMI on ADHD across MR estimators. However, such a finding may be attributed to the more powerful genetic instruments available for BMI compared to ADHD. For example, when including additional genetic variants for ADHD (i.e. using a more liberal threshold when selecting SNPs as instrumental variables), we detected causal effects of ADHD on BMI that were undetected in a previous MR report (23). A more powerful GWAS for ADHD may, in the future, result in consistent findings for the effect of ADHD liability on BMI.

#### Causal or confounded?

The results from our three genetically informed methods were consistent in highlighting multiple sources of confounding affecting the relationships between ADHD and BMI. First, the fact that there was little evidence of an effect in MZ twin-differences analyses suggests that shared genetic and environmental confounding may account for the relationship between ADHD symptoms and BMI. This finding corroborates previous studies using family-based approaches, showing that a substantial proportion of the association between ADHD and obesity is explained by genetic and environmental confounding (42) (14). However, twin differences analyses can be biased by non-shared factors such as measurement errors, which may lead to attenuations of within-twin pair associations and lower statistical power (43).

Although we did not detect associations between twin differences scores in BMI and twin differences scores in ADHD symptoms, substantial auto-regressive pathways were found for both traits (e.g. twin differences in ADHD symptoms at ages 8 and 12 years). As such, twin differences scores showed some reliability and stability over time, suggesting that any undetected true associations between twin differences scores in ADHD symptoms and BMI were likely small. As such, replication of our twin-differences findings in a larger twin sample is needed to verify whether small bidirectional causal effects between ADHD symptoms and BMI can be excluded. Furthermore, cross-lagged models examine causal relationships at specific time lags (i.e. intervals between data collection points). It is possible that the effects between ADHD symptoms and BMI occurred within a different time frame (e.g. immediate effects or very long-term effects), which went undetected in our designs. Of note is that genetic instruments used in polygenic scores and MR reflect long-term exposures (e.g. long-term increased liability to BMI and ADHD), which may be one important source of divergence between findings from these two methods with findings from time-sensitive cross-lagged analyses.

Second, corroborating previous studies, we identified associations between the polygenic score for BMI and ADHD symptoms, and the polygenic score for ADHD and BMI (18, 44). However, the associations may arise from direct causal effects but also from confounding. Indeed, the within-family polygenic score estimate, which controlled for sources of confounding shared between siblings (e.g. population stratification, dynastic effects and assortative mating) revealed that the associations between the ADHD PS and the BMI PS with their opposite phenotypes were to some extent biased by the aforementioned factors. Furthermore, noticeable attenuation of the between-family estimates after controlling for parental education suggests that dynastic effects related to parental education may constitute an important source of confounding. This is in line with previous studies showing substantial confounding due to familial and parental factors in polygenic score studies of educational attainment (45, 46) and cognitive abilities (37). Our study demonstrates that such confounding is not specific to cognitive traits but may also extend to other traits and outcomes, namely ADHD and BMI.

Third, multivariable MR analyses further implicated confounding in the relationship between ADHD and BMI. We found that the effects between ADHD and BMI were substantially attenuated after including educational attainment in multivariable MR analyses, suggesting that some of the effects between the two phenotypes may be due to educational attainment. These findings correspond to current knowledge that population stratification, assortative mating and dynastic effects can result in biased MR estimates (24). Further work implementing a within-family MR design that accounts for such confounding may provide more robust estimates of the causal effects of BMI on ADHD.

#### The developmental feature of bidirectional relationships between ADHD and BMI

As part of twin-differences and polygenic score analyses, we tested whether effects between ADHD and BMI changed from childhood to adolescence. Notably, the BMI PS had a stronger effect on ADHD symptoms in childhood than in adolescence, suggesting that the genetic liability to higher BMI had stronger influences on ADHD symptoms during the childhood years. Interestingly, a recent study has also identified that higher BMI (47)and obesity may be associated with abnormal development of prefrontal cortex and diminished executive function in children (10).

In the other direction, we found that the association between the ADHD PS and BMI increased from childhood to adolescence, suggesting that ADHD may more likely result in higher BMI in adolescence than in childhood. One plausible explanation for the increasing effects of ADHD with age is that individuals with ADHD symptoms may present more impulsive eating and difficulty in planning regular meals or maintaining a healthy lifestyle (48). Consequently, these behaviours may lead to weight gain in adolescence and beyond, when parental monitoring is less pronounced, and individuals are more autonomous in their food consumption patterns.

## Limitations

In addition to the aforementioned methodological limitations associated with each design and limitations related to power, the following limitations should be considered. As we used BMI as the studied variable for our analyses, the interpretation may not be applicable when using obesity as the target. Similarly, ADHD symptoms were dimensionally assessed in our study sample, so the findings may not be readily applicable to clinical samples.

The GWAS for ADHD used in the current study is based on binary diagnosis. However, it is strongly concordant with the GWAS on childhood ADHD symptoms (genetic correlation=0.97) (15). In addition, previous studies have also demonstrated that the polygenic scores generated from the GWAS on ADHD diagnosis were highly correlated with dimensional ADHD symptoms in the general population (49) (50). Such high correlation suggests that risk alleles for the categorical diagnosis of ADHD overlap with those for inattentive and hyperactive/impulsive symptoms dimensionally distributed in the population. Finally, as we used ADHD liability as the exposure variable in MR analyses to study the effect of ADHD on BMI, this may potentially lead to violation of MR assumptions if there is a continuous effect of ADHD on BMI and hence prevents us from estimating the actual effect (51).

## Conclusion

The three genetically informed methods implemented in this study converged to demonstrate that the relationships between ADHD and BMI are subject to shared genetic and environmental confounding. Polygenic score and MR analyses suggest plausible reciprocal causal relationships while findings from cross-lagged analyses were inconsistent. Polygenic score analyses further suggested that reciprocal relationships may be age specific. Future research using larger samples and additional designs are required to provide a definitive answer. A developmentally sensitive approach aiming to describe the timing of the causal effects may be required to elucidate apparently diverging findings. Such a developmentally sensitive approach will also provide invaluable information for prevention and early intervention programmes.

## Data Availability

Summary statistics of the GWAS are publicly available and the TEDS data are available via data request.

https://www.med.unc.edu/pgc/results-and-downloads

https://zenodo.org/record/1251813#.Xph97C2ZNTZ

https://www.ted.ac.uk.

## Acknowledgements

The authors gratefully acknowledge the ongoing contribution of the participants in the Twins Early Development Study and their families.

## Declaration of Conflicting Interests

The authors declared no potential conflicts of interest with respect to the research, authorship and publication of this article.

## Funding

The author(s) disclosed receipt of the following financial support for the research, authorship, and/or publication of this article: The Twins Early Development Study is supported by grant MR/M021475/1 (and previously G0901245) from the UK Medical Research Council. J.-B.P. JBP is a fellow of MQ: Transforming Mental Health (MQ16IP16) and is supported by the Medical Research Foundation 2018 Emerging Leaders 1st Prize in Adolescent Mental Health. C.-Y.L. is supported by an Overseas Research Scholarship from University College London. The Medical Research Council (MRC) and the University of Bristol support the MRC Integrative Epidemiology Unit [MC_UU_00011/1]. NMD is supported by a Norwegian Research Council Grant number 295989.

## Notes

### Competing Interest Statement

The authors have declared no competing interest.

### Summary of Updates

Manuscript, tables and figures have been updated.

